# Associations Between Vestibular Function and White Matter Microstructure in Healthy Older Adults

**DOI:** 10.1101/2025.09.04.25335115

**Authors:** Dominic Padova, Yuchen Yang, J. Tilak Ratnanather, Andreia V. Faria, Yuri Agrawal

**Affiliations:** Center for Imaging Science, Department of Biomedical Engineering, Johns Hopkins University, Baltimore, MD, USA; Li Ka Shing Faculty of Medicine, The University of Hong, Kong Hong Kong, China; Center for Imaging Science and Institute for Computational Medicine, Department of Biomedical Engineering, Johns Hopkins University, Baltimore, MD, USA; Department of Radiology, Johns Hopkins University School of Medicine, Baltimore, MD, USA; Department of Otolaryngology – Head and Neck Surgery, Johns Hopkins University School of Medicine, Baltimore, MD, USA; Department of Otolaryngology – Head and Neck Surgery, University of Colorado Anschutz Medical Campus, Aurora, CO, USA

**Keywords:** Aging, Vestibular, VEMP, VOR, DTI, White Matter

## Abstract

The relationship between vestibular function and white matter (WM) integrity is poorly understood, despite increasing evidence linking vestibular sensory decline and motor and cognitive deficits in older adults. This study examined associations between vestibular function and the microstructural integrity of 19 WM pathways in 394 cognitively healthy participants aged 60+ from the Baltimore Longitudinal Study of Aging who had regional WM microstructure assessed by diffusion tensor imaging metrics: fractional anisotropy and mean diffusivity. The vestibular functions of the saccule, utricle, and horizontal semi-circular canal were assessed using the cVEMP and oVEMP tests and the vHIT, respectively. Multiple linear regression of WM metrics onto vestibular function, adjusted for age, sex, intracranial volume, and scanner type, was used to examine the association between WM and vestibular function. We found that higher canal function was associated with higher integrity in the external capsule, the cingulum projection to the hippocampus, the inferior fronto-occipital fasciculus, and the genu of the corpus callosum, but lower integrity in the limbs of the internal capsule, sagittal stratum, posterior thalamic radiation, the cingulum projection to the cingulate gyrus, and fornix. Utricular function was associated with higher integrity in the anterior limb of the internal capsule and the fornix but lower integrity in the retrolenticular limb of the internal capsule. Saccular function was not associated with WM microstructure. This study demonstrates for the first time potential associations between vestibular end-organ function and WM microstructure in healthy, older adults, with horizontal canal function exhibiting diffuse associations with multi-sensorimotor tracts, whereas utricular function shows more focal associations. Because these findings are concomitant with lower age-related WM integrity, they suggest that better vestibular function may confer resilience in certain pathways while potentially accelerating age-related degeneration in others. Longitudinal studies will be needed to robustly identify WM neuroimaging markers of aging-associated vestibular loss over time.

## 1 Introduction

The vestibular system senses six self-motion cues relative to 3D space and gravity: the two otoliths, the utricle and the saccule, sense 3D translations the orientation with respect to gravity, and the three semi-circular canals sense 3D rotations. This vestibular information is integrated with sensory inputs such as touch, vision, hearing, and proprioception, and is widely distributed throughout the brain via white matter (WM) networks. These networks support motor functions, including balance, posture, and gaze stabilization, as well as self-motion perception during locomotion [1, 2, 3, 4, 5]. Additionally, a plethora of evidence suggests that vestibular information is involved in higher-order cognitive processes, including visuospatial memory and navigation, executive function, attention, motor planning, affective regulation, and bodily self-consciousness [6, 7, 8, 9, 10, 11, 12, 13, 14].

Vestibular structure and function decline with aging [15]. Such declines have been linked to gray matter alterations in a diverse array of brain regions and to deficits in these motor and cognitive domains, even in the absence of overt clinical symptoms [16, 17, 18, 19, 20, 21, 9]. Furthermore, vestibular dysfunction, gray matter alterations in the vestibular network, and impairments in motor and cognitive abilities are common features of several neurodegenerative diseases, such as Alzheimer’s disease, multiple sclerosis, Huntington’s disease, and Parkinson’s disease which progress with aging and involve the degradation of WM networks [22, 23, 24, 25, 26, 27, 28, 29, 30, 31, 32, 33, 34, 35, 36, 37]. However, the alterations in WM microstructure related to age-related vestibular dysfunction are poorly characterized.

Network-based research on vestibular dysfunction suggests a reorganization of the functional connectivity between myriad brain regions that depends on etiology [38, 39, 40]. Because functional networks depend on the WM that connects gray matter regions, it is therefore crucial to understand the spatial patterns of WM alterations in age-related vestibular dysfunction. Diffusion tensor imaging (DTI) is an MRI technique that has been used to sensitively detect microstructural changes in WM in aging that are related to cognitive decline [41, 42]. Several DTI studies have demonstrated that damage to vestibular WM networks is associated with impairments in self-motion perception, graviception, balance, posture, and gait [43, 44, 45, 46, 47, 35]. Additionally, DTI studies revealed that age-related declines in peripheral vestibular function and WM integrity have been linked to balance, posture, and dual-task gait impairments and to increased risk of neurodegenerative disorders, such as Alzheimer’s disease [48, 49, 50, 51, 52, 53, 22, 25, 54]. Despite these connections, limited research has examined the relationship between age-related vestibular function and WM microstructure.

The objective of this cross-sectional study of 394 healthy older adults is to examine the association between age-related variation in vestibular end-organ function and microstructural integrity in 19 WM tracts involved in higher order behaviors, while accounting for age, intracranial volume, sex, and scanner protocol. These tracts include the cingulum (hippocampus, cingulate gyrus), external capsule, limbs of the internal capsule (anterior, posterior, retrolenticular), cerebral peduncle, posterior thalamic radiation, corona radiata (anterior, posterior, superior), fronto-occipital fasciculus (inferior, superior), superior longitudinal fasciculus, uncinate fasciculus, sagittal stratum, and corpus callosum (genu, body, splenium). We hypothesized that better age-related vestibular function will differentially correlate with higher WM integrity, analogously to prior research on vestibular function and balance. To ensure that vascular risk or multisensory function do not confound or obscure the effects of vestibular function, we perform follow-up analyses additionally covarying for hyperlipidemia, hypertension, diabetes, smoking status, and sensory functions (visual acuity, proprioception, and hearing).

## 2 Data and methods

### 2.1 Study sample

Our study sample includes 394 participants aged 60+ from the Baltimore Longitudinal Study of Aging (BLSA) [55]. All participants were cognitively normal and had no diagnosis of stroke, head injuries, or neurodegenerative disease. All subjects gave written informed consent. The study was approved by the Internal Review Board of the Intramural Research Program of the National Institutes of Health.

Having diabetes, smoking status, hypertension, hearing loss, visual acuity loss, and proprioceptive loss were measured and included as confounding variables in follow-up hypothesis tests. Participants were classified as diabetic if a doctor had told them they had diabetes, as a smoker if they had responded affirmatively to have ever smoked 100 cigarettes, sever smoked 50 cigars, or ever smoked 3 packages pipe tobacco, as hyperlipidemic if they had high blood cholesterol, and as hypertensive if a doctor had ever told them that they were hypertensive. Hearing loss was measured as the speech-frequency pure tone average of air-conduction thresholds at 0.5, 1, 2, and 4 kHz from the better ear. Visual acuity loss, which refers to how much a pattern must differ in size to be seen, was measured as the angular deviation in logMAR units and ranges from 0.80 to -0.30 logMAR, where lower values indicate better acuity. Proprioceptive loss was measured as the degree of ankle deflection perceptible according to an established BLSA procedure [56]. To maintain the interpretation that higher values mean better function, the hearing, vision, and proprioceptive variables were treated as continuous variables and were negated.

### 2.2 Vestibular physiologic testing

Following established procedures, the functions of the saccule, utricle, and horizontal semicircular canal were assessed using the cervical vestibular evoked myogenic potential (cVEMP) test, ocular VEMP (oVEMP) test, and video head-impulse test (vHIT), respectively, as described below [22, 57, 58, 59]. For the statistical analysis, continuous vestibular variables were z-score transformed, and categorical variables were created based on whether a response was present on any side for cVEMP and oVEMP, and whether the reported mean VOR gain from the vHIT was below 0.8. To record cVEMP and oVEMP, a commercial electromyographic system (software version 14.1, Carefusion Synergy, Dublin, OH) was used. Electromyogram (EMG) signals were recorded with disposable, pre-gelled Ag/AgCl electrodes with 40-inch safety lead wires from GN Otometrics (Schaumburg, IL). EMG signals were amplified and band-pass filtered using 20–2000 Hz for cVEMP and 3–500 Hz for oVEMP.

#### 2.2.1 Cervical vestibular-evoked myogenic potential (cVEMP) test

The cVEMP test measures the function of the saccule (and inferior vestibular nerve) [22, 57, 58, 59]. Participants sat on a chair inclined at 30° above the horizontal plane. Trained examiners positioned EMG electrodes bilaterally on the sternocleidomastoid and sternoclavicular junction, with a ground electrode on the manubrium sterni. Participants were instructed to turn their heads to generate at least a 30 *µ*V background response prior to delivering sound stimuli. Bursts of 100 auditory stimuli stimuli of 500 Hz and 125 dB were administered monoaurally through headphones (VIASYS Healthcare, Madison, WI). cVEMPs were recorded as short-latency EMGs of the inhibitory response of the ipsilateral sternocleidomastoid muscle. To calculate corrected cVEMP amplitudes, nuisance background EMG activity collected 10 ms prior to the onset of the auditory stimulus were removed. The higher corrected cVEMP amplitude (unitless) from the left and right sides was used as a continuous measure of saccular function. A difference of 0.5 in corrected cVEMP is considered clinically relevant [57]. An absent response was defined by a response below a threshold level per published guidelines. If this occurred, the assessment was repeated to confirm an absent response. For participants with a present response, the corrected cVEMP amplitude of the better ear was used in the analysis.

#### 2.2.2. Ocular vestibular-evoked myogenic potential (oVEMP) test

The oVEMP test measures the function of the utricle (and superior vestibular nerve) [22, 57, 58, 59]. Participants sat on a chair inclined at 30° above the horizontal plane. Trained examiners placed a noninverting electrode ≈3 mm below the eye centered below the pupil, an inverting electrode 2 cm below the noninverting electrode, and a ground electrode on the manubrium sterni. To ensure that symmetric signals are recorded from both eyes, participants were instructed to perform multiple 20° vertical saccades before stimulation. During oVEMP testing, participants were instructed to maintain an upward gaze of 20°. Head taps (vibration stimuli) applied to the midline of the face at the hairline and ≈30% of the distance between the inion and nasion using a reflex hammer (Aesculap model ACO12C, Center Valley, PA). oVEMPs were recorded as short-latency EMGs of the excitation response of the contralateral external oblique muscle of the eye. The higher oVEMP amplitude (*µ*V) from the left and right sides was used as a continuous measure of utricular function. A difference of 5 *µ*V in oVEMP is considered clinically relevant [57]. If the response was below threshold levels, an absent response was recorded. If this occurred, the assessment was repeated to confirm an absent response. For participants with a present response, the oVEMP amplitude of the better ear was used for analysis.

#### 2.2.3 Video head impulse test (vHIT)

The vHIT measures the horizontal vestibular-ocular reflex (VOR) [22, 60, 61] and was performed using the EyeSeeCam system (Interacoustics, Eden Prarie, MN) in the same plane as the right and left horizontal semicircular canals [61, 62, 63]. To position the horizontal canals in the plane of stimulation, trained examiners tilted the participant’s head downward 30° below the horizontal plane and instructed participants to maintain their gaze on a wall target ≈1.5 m away. The examiner delivered rotations of 5-10° (≈150-250° per second) to the participant’s head. The head impulses are performed at least 10 times parallel to the ground toward the right and left, chosen randomly so that the participant could not predict the direction. The EyeSeeCam system quantified eye and head velocity. VOR gain was calculated as the unitless ratio of the eye velocity to the head velocity. A VOR gain equal to 1.0 is normal and indicates equal eye and head velocities. The mean VOR gain from the left and right sides was used as a continuous variable. A difference of 0.1 in VOR gain is considered clinically relevant [22, 57]. Having a VOR gain less than 0.8, indicating peripheral vestibular hypofunction, was used as a categorical variable.

### 2.3 Diffusion tensor imaging acquisition

Diffusion-weighted MRI scans were acquired on three, 3.0 Tesla Philips Achieva scanners. For scanners 1 and 2, the DTI protocol was the same (acquired at the Kennedy Krieger Institute; number of gradients: 32, number of B0 images = 1, max b-factor = 700 s/mm2, TR/TE = 6801/71 ms, number of slices = 65, voxel size = 0.83 x 0.83 x 2.2 mm, reconstruction matrix = 256 x 256, acquisition matrix = 96 x 95, field of view = 212 x 212 mm, flip angle = 90°), and differed from the scanner 3 protocol (acquired at the National Institute on Aging; number of gradients: 32, number of B0 images = 1, max b-factor = 700 s/mm2, TR/TE = 7454/75 ms, number of slices = 70, voxel size = 0.81 x 0.81 x 2.2 mm, reconstruction matrix = 320 x 320, acquisition matrix = 116 x 115, field of view = 260 x 260 mm, flip angle = 90°). Two B0 images averaged in k-space were included for each DTI acquisition. Previous work has examined the reliability of diffusion measures across scanners in the BLSA and found acceptable levels of test–retest reliability [64].

### 2.4 Diffusion tensor imaging processing pipeline

DTI processing followed previously established procedures for tensor fitting and quality control [65, 66]. Individual diffusion-weighted images were affinely registered to B0 images to correct for physiological motion effects and eddy currents using Functional Magnetic Resonance Imaging of the Brain Software Library (FSL) eddy_correct tool. The rotational component was corrected in the gradient tables using finite strain [67]. Prior to tensor fitting, each diffusion-weighted image was normalized by its own B0 image to combine the two DTI sessions that had different unknown intensity normalization constants. To enhance the robustness of the voxel-wise tensor estimation procedure, iteratively reweighted least squares fitting with outlier rejection was used in the form of RESTORE [68] from the Camino toolkit [69]. Quality control included removing scans with visually identified artifacts, such as excessive motion, corrupted scans, or globally high diffusion measure bias, in which outliers were defined as having >2 standard deviations. [65, 66]. Quality assurance review flagged 50 sessions for removal from further analyses, yielding 394 good quality DTI sessions for analysis. The resultant mean diffusivity (MD) and fractional anisotropy (FA) maps from good quality tensor fitting were selected for analysis.

Image segmentation followed previously established procedures [70]. Briefly, the Johns Hopkins University (JHU) MNI-SS Atlas (Single Subject) [71] was non-rigidly registered to the target scans using Advanced Normalization Tool Symmetric Image Normalization (ANTs SyN). To segment the images, the JHU MNI-SS WM labels were combined with corresponding WM labels from a multi-atlas segmentation using 35 manually labeled atlases from NeuroMorphometrics with the BrainCOLOR protocol [72] and FA-mapped MRI. To remove registration errors of the single-atlas approach, the JHU MNI-SS WM labels were intersected with the WM segmentation from BrainCOLOR. Then the intersected labels were iteratively dilated to fill the remaining WM space from the multi-atlas labels. Then the WM labels obtained from the T1-weighted image were affinely registered to the FA and MD images and used to extract average FA and MD values for each label.

The WM tracts in this study were selected *a priori* based on their known associations with higher-order abilities, such as mobility, attention, and executive, spatial, and affective functions. Thus, WM tract integrity of 19 WM regions pertaining to seven WM tract pathways (illustrated in Figure 1) was assessed. The WM of the fornix (cres), stria terminalis, and tapetum were excluded from the analysis because they could not be reliably resolved with the current imaging resolution. All WM tract regions were considered unilaterally except regions of the corpus callosum due to its nature of bilateral information transmission. Ordinarily, higher WM tract integrity is indicated by higher FA and lower MD. However, in order to ease the interpretation of the MD measure so that higher values indicate higher WM integrity, we have multiplied the MD values by *−*1 and called this new variable -MD.

**Figure 1:**
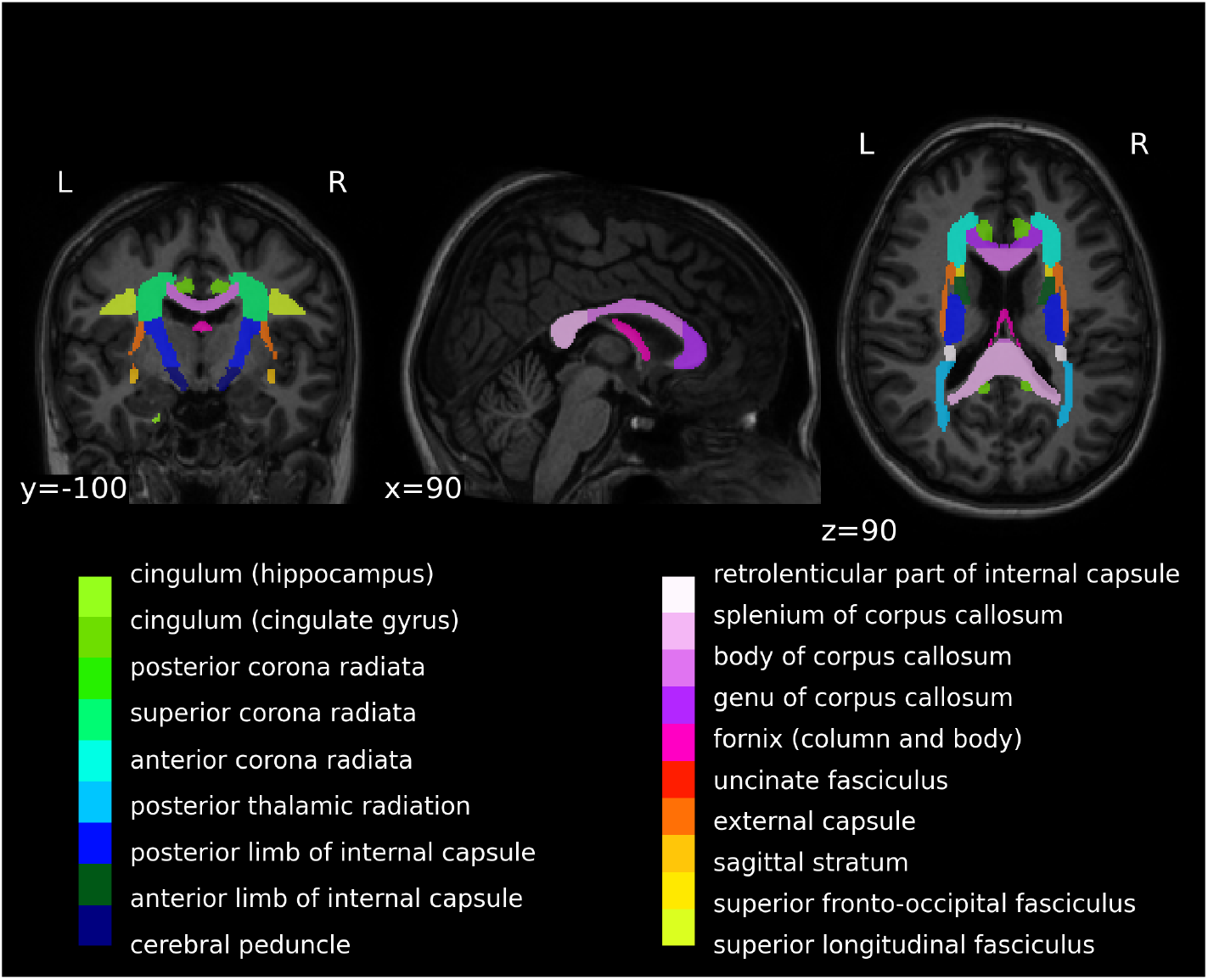
JHU MNI-SS Selected ROIs. Coronal, sagittal, and axial slices showing the JHU MNI-SS WM ROIs overlaid on the T1-weighted image [71].

### 2.5 Statistical modeling

Standardized multiple linear regression was used to assess the relationship between WM tract integrity and vestibular end-organ function, correcting for age, sex, intracranial volume, and scanner protocol. Eq. (1) shows the null hypothesis (*H*_0_) based on which a DTI metric of participant *i* ∈ {1, 2, *…, N, DTI*_*i*_} is predicted. Eq. (2) is the alternative hypothesis (*H*_1_) predicting a DTI metric but now using a vestibular testing variable *vest*_*i*_, both continuous and categorical (coded as 0 for impaired or 1 unimpaired),

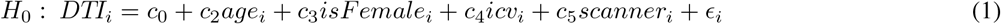

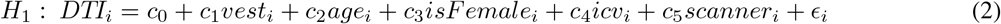

where *age*_*i*_ is the age of subject *i, sex*_*i*_ is the binary categorical variable of subject *i* (female = 1, male = 0), *icv*_*i*_ is the intracranial volume of subject *i*, and *scanner*_*i*_ is the DTI scanner protocol used for subject *i*. All continuous variables were standardized before regression to have a mean of zero and standard deviation of one. The set of unknown coefficient 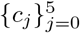 were estimated using maximum likelihood. To robustly evaluate the statistical significance of the addition of vestibular function to our models, we permuted model residuals under the null hypothesis (*n*_*perm*_ = 10, 000 simulations) according to Eq. (3), with significance determined by the proportion of simulated likelihood ratio test (LRT) statistics, *LRT*_*sim*_, larger than the observed LRT, *LRT*_*obs*_, falling below the 0.05 significance level:

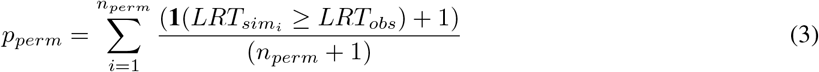

where **1**(·) is the indicator function which outputs a value of 1 when 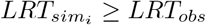 and 0 otherwise. Additional *χ*^2^-tests were performed to test our hypotheses. For models that reject the null hypothesis, *H*_0_, (*p*_*perm*_ *<* 0.05), 95% confidence intervals calculated by bootstrapping model residuals under the alternative hypothesis, *H*_1_, (*n*_*boot*_ = 10, 000 simulations), using custom functions with the *np*.*boot* function from the npboot R package. All reported permutation p-values are approximate, as indicated by “≈” [73]. False discovery rate correction of observed p-values (i.e. p-values for the vestibular terms in the observed model, not *p*_*perm*_) was performed using the Benjamini-Hotchberg method across both DTI metrics and all investigated structures to yield *q*_*all*_. We additionally report less conservative q-values in which observed p-values were corrected across both DTI metrics and the structures of each hemisphere separately yielding *q*_*left*_, *q*_*right*_ as well as across the callosal tracts alone yielding *q*_*callosal*_. Significance was determined at the 0.05 level.

To test whether the addition of the function of hearing, vision, or proprioception either explains away or masked vestibular relationships, we performed follow-up multi-sensory hypothesis tests. The null hypothesis, *H*_0*B*_, in Eq. (4) and the alternative hypothesis, *H*_1*B*_, in Eq. (5) additionally covary for the *hearing, vision, proprioception* variables and their interactions with vestibular function,

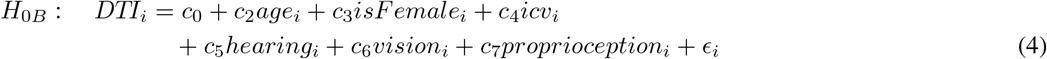

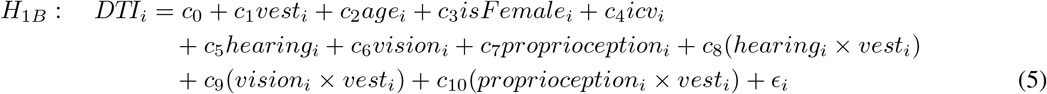

To test our hypothesis, *χ*^2^-tests were performed first to assess model significance, then Wald tests were used to determine the significance of the individual vestibular estimates within the models that rejected the null hypothesis *H*_0*B*_ according to the *χ*^2^-test. A similar analysis was performed for the follow-up test of vascular risk factors *diabetes, hypertensive, smoking, hyperlipidemic* and their interactions with vestibular function. All statistical analyses were performed using RStudio [74].

## 3 Results

### 3.1 Characteristics of the study sample

Table 1 shows the characteristics for the study sample from the BLSA.

**Table 1:** Characteristics of the study sample (n = 394). Key: PTA: four-frequency (0.5, 1, 2, 4 kHz) pure tone average from the better ear; N: the number of participants with a visit where both the characteristic and MRI data were available; %: 100(N/n) percent; SD: standard deviation.

| Characteristic | Mean (SD) | N (%) |
| --- | --- | --- |
| Age (years) | 73.3 (7.97) |  |
| Sex |  |  |
| Male |  | 166 (42.1) |
| Female |  | 228 (57.9) |
| Education (years) | 16.9 (2.39) |  |
| Corrected cVEMP Amplitude | 1.3 (0.977) |  |
| oVEMP Amplitude (uV) | 12.2 (7.74) |  |
| Mean VOR Gain | 0.989 (0.177) |  |
| cVEMP Bilaterally Absent |  |  |
| No (0) |  | 228 (57.9) |
| Yes (1) |  | 135 (34.3) |
| oVEMP Bilaterally Absent |  |  |
| No (0) |  | 280 (71.1) |
| Yes (1) |  | 87 (22.1) |
| VOR Mean Impaired |  |  |
| No (0) |  | 312 (79.2) |
| Yes (1) |  | 46 (11.7) |
| Proprioception Threshold (degrees) | 1.93 (2.03) |  |
| Four Frequency PTA (dB) | 32.4 (15) |  |
| Visual Acuity (logMAR) | 0.116 (0.154) |  |
| Hypertensive |  |  |
| No (0) |  | 198 (50.3) |
| Yes (1) |  | 194 (49.2) |
| Diabetic |  |  |
| No (0) |  | 333 (84.5) |
| Yes (1) |  | 61 (15.5) |
| Hyperlipidemic |  |  |
| No (0) |  | 158 (40.1) |
| Yes (1) |  | 234 (59.4) |
| Smoking History |  |  |
| Never Smoked (0) |  | 214 (54.3) |
| Smoker (1) |  | 180 (45.7) |

### 3.2 Vestibular Effects Found by Linear Regression

For the interpretation of all results, recall that we have recoded the MD, indicated by “-MD”, and the categorical vestibular measures so that higher values mean better integrity. Additionally, all continuous variables were z-scored, including the DTI and continuous vestibular measures. Therefore, the effect sizes indicate the z-score increase in WM integrity for a one z-score increase in vestibular function. Tables 2 and 3 summarize the results of the main and follow-up analyses of the continuous and categorical vestibular variables, respectively. Figures 2 and 3 respectively illustrate the spatial distribution of the significant continuous and categorical vestibular-only effects on -MD and FA from the main alternative hypothesis *H*_1_. All *χ*^2^-tests performed agreed with the outcomes of permutation testing (results not shown).

**Table 2:** Summary of associations with continuous vestibular variables. The p-values in the main analysis results are the permutation p-values, whereas p-values in the follow-up analyses are the Wald p-values. (Abbreviations: DTI = diffusion tensor imaging metric, ALIC = anterior limb of internal capsule, PLIC = posterior limb of internal capsule, RLIC = retrolenticular part of internal capsule, EC = external capsule, SS = sagittal stratum, PTR = posterior thalamic radiation, IFO = inferior fronto-occipital fasciculus, CH = cingulum (hippocampus), GCC = genu of corpus callosum, DM = diabetes mellitus, FA = fractional anisotropy, MD = mean diffusivity, CI = 95% confidence interval, SE = standard error, * = *q*_*all*_ *<* 0.05, *†* = *q*_*left*_ or *q*_*right*_ *<* 0.05.)

| Vestib. | WM Region | DTI | Effect Size | *p* | WM Int. |
| --- | --- | --- | --- | --- | --- |
| Main Analysis |  |  |  |  |  |
| VOR | L ALIC | -MD | -0.097 (CI: (-0.18, -0.016)) | 0.021 | ↓ |
|  | L PLIC | -MD | -0.12 (CI: (-0.20, -0.017)) | 0.022 | ↓ |
|  | L RLIC | -MD | -0.088 (CI: (-0.17, -0.0027)) | 0.042 | ↓ |
|  | L SS | FA | -6.7e-13 (CI: (-1.06e-12, -2.99e-13)) | 0.001*,† | ↓ |
|  | L SS | -MD | -0.093 (CI: (-0.18, -0.0099)) | 0.034 | ↓ |
|  | L PTR | FA | -0.11 (CI: (-0.25, -0.035)) | 0.025 | ↓ |
|  | L IFO | FA | 0.12 (CI: (0.046, 0.26)) | 0.018 | ↑ |
|  | L CH | -MD | 0.1 (CI: (0.018, 0.19)) | 0.022 | ↑ |
|  | L EC | FA | 0.13 (CI: (0.027, 0.3)) | 0.043 | ↑ |
|  | R EC | FA | 0.15 (CI: (0.064, 0.29)) | 0.0098 | ↑ |
|  | GCC | -MD | 0.096 (CI: (0.015, 0.18)) | 0.030 | ↑ |
| oVEMP | R RLIC | FA | -0.18 (CI: (-0.35, 0.0054)) | 0.042*,† | ↓ |
| cVEMP | — | — | No significant correlation |  |  |
| Follow-up (Covarying for Vascular Risk) |  |  |  |  |  |
| VOR | GCC | FA | No significant correlation |  |  |
| Interaction: no hyperlipidemia |  |  | -0.29 (SE: 0.087) | 9.3e-4 | ↓ |
| VOR | L IFO | FA | No significant correlation |  |  |
| Interaction: no hypertension |  |  | -0.3 (SE: 0.092) | 7.8e-4 | ↓ |
| Interaction: no hyperlipidemia |  |  | 0.37 (SE: 0.099) | 2.6e-4 | ↑ |
| Interaction: no smoking |  |  | -0.18 (SE: 0.095) | 0.058 | ↓ (trend) |
| Follow-up (Covarying for Multisensory Function) |  |  |  |  |  |
| VOR | L IFO | FA | 0.17 (SE: 0.058) | 3.4e-3 | ↑ |
| Interaction: hearing |  |  | 0.12 (SE: 0.061) | 0.048 | ↑ |
| VOR | L PLIC | -MD | -0.082 (SE: 0.049) | 0.095 | ↓ (trend) |
| Interaction: proprio. |  |  | -0.14 (SE: 0.046) | 2.7e-3 | ↓ |
| Interaction: hearing |  |  | 0.12 (SE: 0.053) | 0.023 | ↑ |
| VOR | GCC | FA | No significant correlation |  |  |
| Interaction: hearing |  |  | -0.13 (SE: 0.05) | 0.013 | ↓ |

**Table 3:** Summary of associations with categorical vestibular variables. The p-values in the main analysis results are the permutation p-values, whereas p-values in the follow-up analyses are the Wald p-values. (Abbreviations: DTI = diffusion tensor imaging metric, ALIC = anterior limb of internal capsule, PLIC = posterior limb of internal capsule, CGC = cingulum (cingulate gyrus), FX = column/body of the fornix, GCC = genu of corpus callosum, FA = fractional anisotropy, MD = mean diffusivity, CI = 95% confidence interval, SE = standard error.)

| Vestib. Category | WM Region | DTI | Effect Size | *p* | WM Int. |
| --- | --- | --- | --- | --- | --- |
| Main Analysis |  |  |  |  |  |
| Unimpaired canal(VOR ≥ 0.8) | L PLIC | FA | 0.81 (CI: (0.63, 2.2)) | 0.018 | ↑ |
|  | R CGC | FA | 0.57 (CI: (0.43, 2.29)) | 0.037 | ↑ |
|  | R FX | -MD | -0.52 (CI: (-0.85, -0.20)) | 0.001 | ↓ |
|  | GCC | -MD | -0.35 (CI: (-0.72, -0.044)) | 0.043 | ↓ |
| Any present oVEMP | L FX | FA | 0.36 (CI: (0.20, 0.98)) | 0.039 | ↑ |
|  | R ALIC | FA | 0.36 (CI: (0.15, 0.95)) | 0.049 | ↑ |
| Any present cVEMP | – | – | No significant correlation |  |  |
| Follow-up (Covarying for Multisensory Function) |  |  |  |  |  |
| Unimpaired canal | R FX | -MD | 0.45 (SE: 0.16) | 0.0052 | ↑ |
|  | GCC | -MD | 0.35 (SE: 0.18) | 0.044 | ↑ |
| Follow-up (Covarying for Vascular Risk) |  |  |  |  |  |
| Unimpaired canal | L PLIC | FA | No significant correlation |  |  |
| Interaction: no hypertension |  |  | -1.01 (SE: 0.35) | 0.004 | ↓ |
| Interaction: no smoking |  |  | 1.4 (SE: 0.35) | 8.73e-05 | ↑ |
| Unimpaired canal | R CGC | FA | No significant correlation |  |  |
| Interaction: no hypertension |  |  | 0.73 (SE: 0.36) | 0.041 | ↑ |
| Interaction: no DM |  |  | -0.86 (SE: 0.52) | 0.098 | ↓ (trend) |
| Interaction: no smoking |  |  | 0.73 (SE: 0.36) | 0.046 | ↑ |
| Unimpaired canal | GCC | -MD | 0.73 (SE: 0.41) | 0.078 | ↑ (trend) |
| Interaction: no smoking |  |  | -0.71 (SE: 0.27) | 9.8e-3 | ↓ |

**Figure 2:**
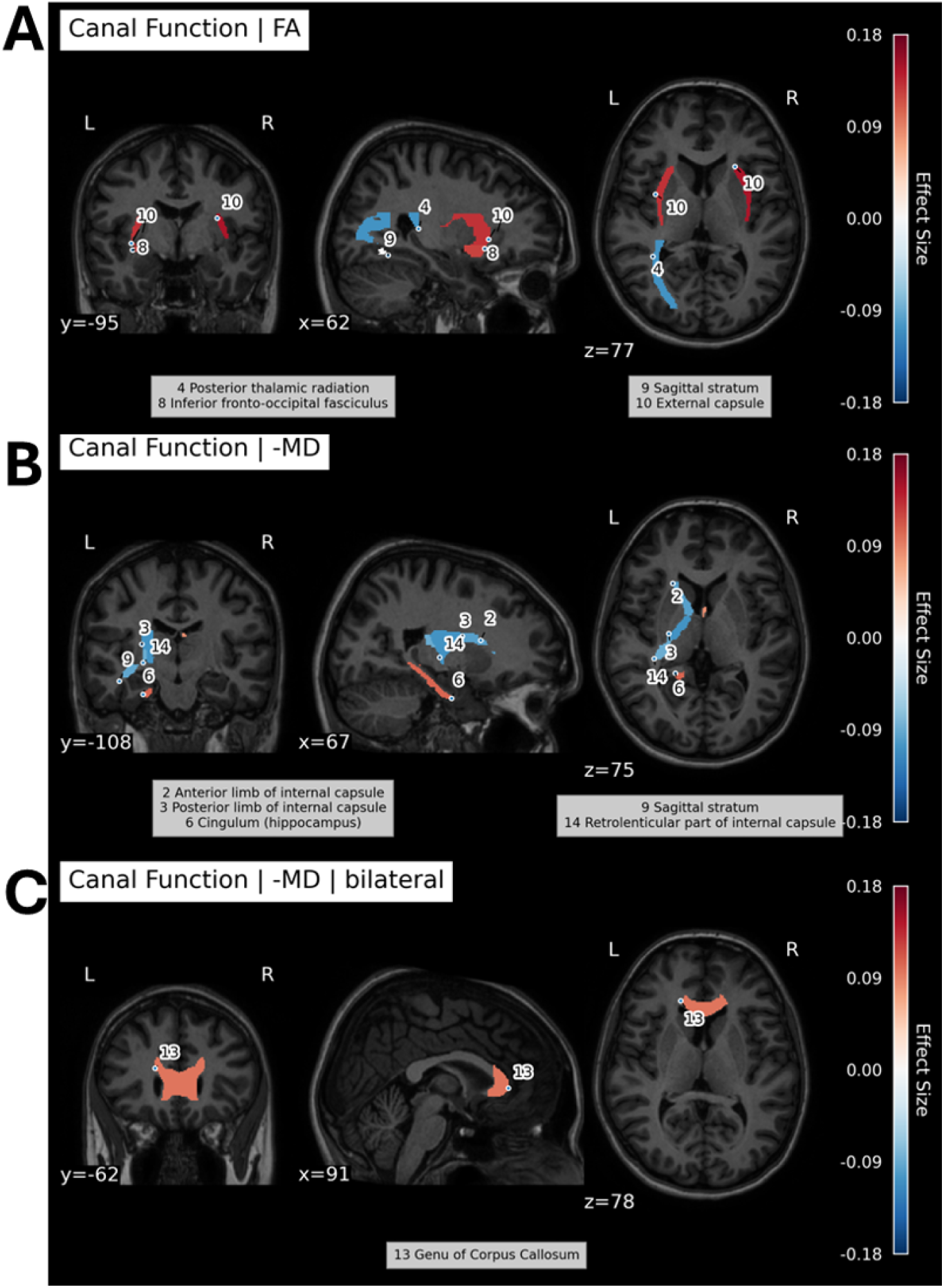
Spatial distribution of the significant horizontal canal effects on lateralized FA (A), on lateralized -MD (B), and on the interhemispheric -MD (C) models visualized on the JHU MNI-SS T1-weighted template.

**Figure 3:**
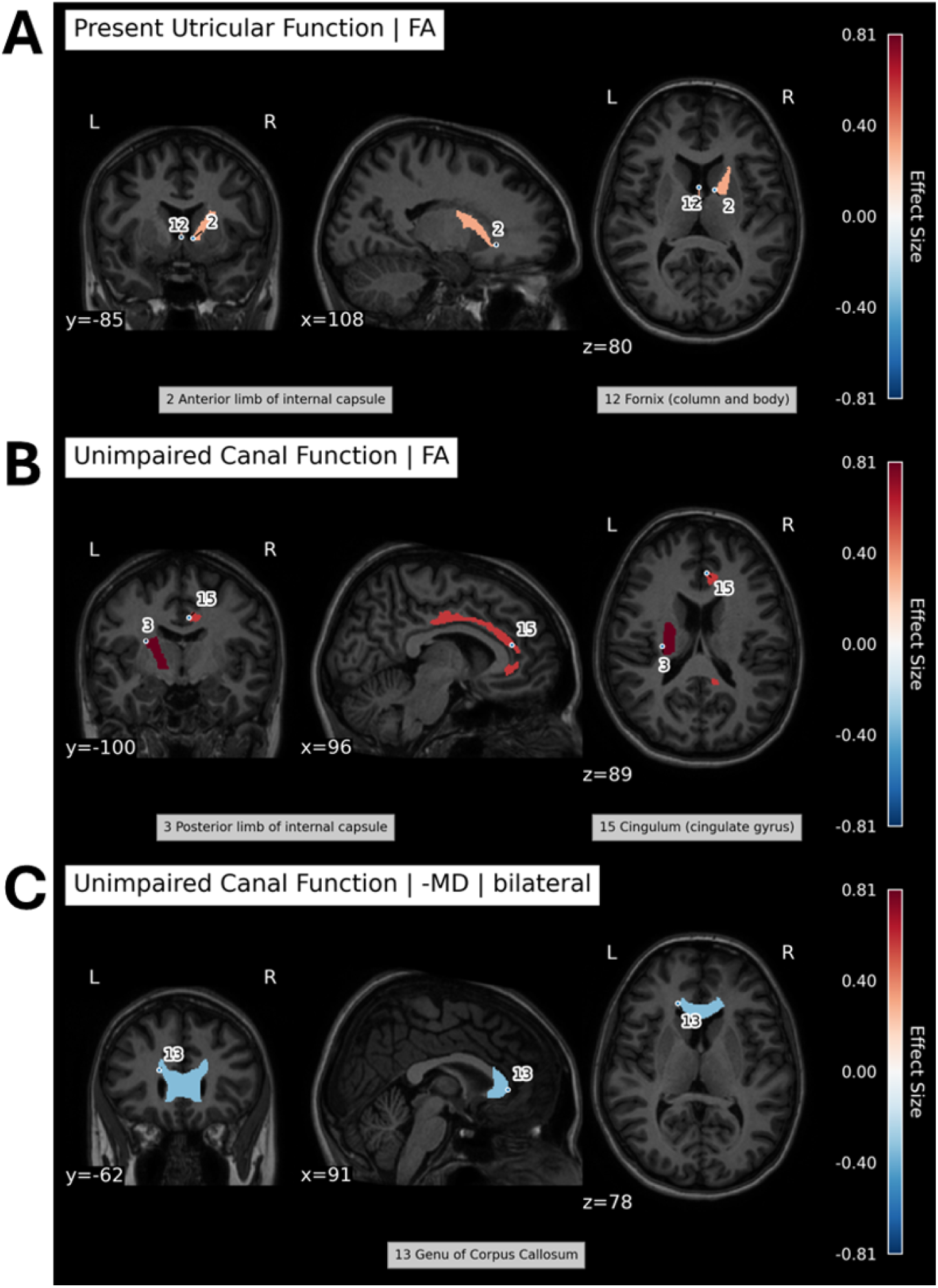
**Spatial distribution of the significant categorical utricular effects on lateralized FA (A) categorical horizontal canal effects on lateralized FA (B) and on the interhemispheric -MD (C) models visualized on the JHU MNI-SS T1-weighted template**.

#### 3.2.1 Continuous Vestibular Variables

In the main analysis (Table 2), higher mean VOR gain (i.e. higher horizontal canal function) was related to poorer WM integrity (lower -MD or lower FA) in several left-hemisphere regions according to permutation testing, including the anterior limb of the internal capsule, the posterior limb of the internal capsule, the retrolenticular part of the internal capsule, the sagittal stratum, and the posterior thalamic radiation. showed a complex relationship with the microstructure of numerous WM regions. By contrast, higher mean VOR gain was association with higher WM integrity (higher -MD or higher FA) in the left inferior fronto-occipital fasciculus, left cingulum (hippocampus), left and right external capsules, and the genu of the corpus callosum. Regarding age-related otolith function, higher oVEMP amplitude was associated with poorer WM integrity (lower FA) in the right retrolenticular part of the internal capsule, whereas cVEMP showed no significant relationships according to permutation testing. Two relationships between continuous vestibular function and WM integrity measures that survived permutation testing also survived FDR-correction at the 0.05 level: utricular function and WM integrity of the right posterior limb of the internal capsule (*q*_*all*_ = 0.023, *q*_*right*_ = 0.021) and canal function and WM integrity of the left sagittal stratum (*q*_*all*_ = 0.058, *q*_*left*_ = 0.029). However, no relationships between continuous vestibular function and WM integrity measures in the corpus callosum that survived permutation testing also survived FDR-correction at the 0.05 level (*q*_*callosal*_ *>* 0.05).

Several results from the primary analysis persisted in the follow-up analyses, while others did not (Table 2). In the follow-up analysis covarying for vascular risk factors, the main effects of mean VOR gain for the genu of the corpus callosum and the left inferior fronto-occipital fasciculus remained significant, whereas the other main effects fell below the significance threshold. Significant interactions indicated that the absence of hyperlipidemia was associated with higher WM integrity in the genu of the corpus callosum, in contrast to its effect in the inferior-fronto-occipital fasciculus. Furthermore, the absence of hypertension was associated with either lower WM integrity in the inferior-fronto-occipital fasciculus. The interaction with smoking status (i.e. no smoking) trended toward an association with lower WM integrity in that tract. In the follow-up analysis covarying for multisensory function, higher VOR gain remained associated with higher WM integrity (higher FA) in the left inferior fronto-occipital fasciculus, along with an interaction with hearing function. Interactions with hearing and proprioception functions indicated that the effect of VOR gain on WM integrity in the left posterior limb of the internal capsule could be positively (lower -MD) and negatively (higher -MD) modified by these additional sensory factors, respectively. No main effect was observed for VOR gain in the genu of the corpus callosum in these models, although an interaction with hearing suggested lower WM integrity (lower FA) in the presence of better VOR gain and better hearing function.

#### 3.2.2 Categorical Vestibular Variables

Table 3 shows the main analysis of categorical vestibular function and WM integrity. Having unimpaired canal function (i.e., a mean VOR gain ≥ 0.8) was correlated with higher WM integrity (higher FA or lower -MD) of the left posterior limb of the internal capsule, the right cingulum (cingulate gyrus), the right column and body of the fornix, and the right anterior limb of the internal capsule. Compared to participants with bilaterally absent oVEMP responses, having any present oVEMP response was correlated with higher WM integrity (higher FA) of the left column and body of the fornix and the right anterior limb of the internal capsule. Having any present cVEMP response was not correlated with WM integrity as measured by FA or MD. No relationships between categorical vestibular function and WM integrity measures that survived permutation testing also survived FDR-correction at the 0.05 level (*q*_*all*_, *q*_*left*_, *q*_*callosal*_ *>* 0.05). However, one relationship showed a strong trend: unimpaired canal function and WM integrity of the right column and body of the fornix (*q*_*right*_ = 0.059).

In the follow-up analysis covarying for vascular risk factors, having unimpaired canal function showed no main association with WM integrity in the left posterior limb of the internal capsule or in the right cingulum (cingulate gyrus). However, interaction effects emerged with specific vascular risk factors. In the left posterior limb of the internal capsule, the absence of hypertension was related to lower FA, whereas no smoking history was linked to higher FA. In the right cingulum, both the absence of hypertension and no smoking history were associated with higher FA, whereas not having diabetes mellitus showed a trend toward lower FA. Additionally, unimpaired canal function showed a strong trend toward association with higher WM integrity (higher -MD) in the genu of the corpus callosum, and the interaction of smoking status (i.e. no history of smoking) was associated with lower WM integrity (lower -MD). In the follow-up analysis covarying for multisensory function, having unimpaired canal function remained associated with higher WM integrity (higher -MD) in the right column and body of the fornix and the genu of the corpus callosum (Table 3).

## 4 Discussion

This study is one of the first to demonstrate that age-related variation in vestibular function may be differentially associated with the structural integrity of multiple WM tracts thought to transmit vestibular and multi-sensorimotor information to support motor and cognitive abilities. Our analysis revealed that horizontal canal function was the most associated with WM microstructure, and that the most frequently occurring WM changes associated with vestibular function were found in the limbs of the internal capsule, the cingulum, the left fronto-occipital fasciculus, and the genu of the corpus callosum. These affected regions overlap with those found in a previous study linking chronic bilateral vestibulopathy with reduced FA (poorer WM integrity) [75]. However, these results were affected by adjustment for vascular risk factors and multisensory function. Once vascular risk factors were added to the models, some of the vestibular effects were no longer present as main effects, and instead emerged as significant interaction effects with specific risk factors (e.g. no hyperlipidemia, no hypertension, no smoking, no diabetes). This suggests that vascular health may modulate the link between vestibular function and WM microstructure. In the models controlling for multisensory function, few main vestibular associations persisted and several interactions with hearing or proprioception emerged. The positive (negative) interactions indicate that, as hearing/proprioception function increases, the effect of vestibular function on WM integrity becomes more positive/less negative (more negative/less positive). In certain WM pathways (e.g., left inferior fronto-occipital fasciculus, left posterior limb of the internal capsule, genu of the corpus callosum), higher vestibular function was more strongly linked to higher WM integrity with higher hearing or proprioception function. This suggests a multisensory compensation or synergy that may be pathway-specific.

Although only two relationships survived FDR-correction due to the large number of comparisons, numerous interesting trends emerged at an uncorrected threshold and merit further investigation and discussion. In the following sections, we first consider how aging of peripheral vestibular function intersects with changes in white matter structure (Section 4.1). We then discuss how these changes relate to higher-order behaviors, specifically focusing on sensorimotor control (Section 4.2.1) and cognitive-sensorimotor interactions (Section 4.2.2).

### 4.1 Aging of Vestibular Function and WM Structure

These findings complement the growing body of literature linking aging and vestibular function to WM structure. The number of vestibular ganglion cells and hair cells decline with aging, with the rate of hair cell decline in the cristae of the semi-circular canals being higher than that of the otolithic maculae [76, 77, 78, 79]. The accumulated vestibular damage leads to different rates of (linear) declines of otolith and canal functions [15]. Despite low levels of vestibular hair cell regeneration and central vestibular compensation, such age-related declines are associated with motor deficits, cognitive risk, and future risk of falls and neurodegenerative disorders that impact gray and WM, such as Alzheimer’s disease [48, 49, 50, 51, 52, 53, 22, 25, 54]. The exact mechanisms by which age-related vestibular loss may impact WM microstructure are poorly characterized. One possible mechanism is that peripheral vestibular sensory loss leads to afferent neuron degeneration, which in turn leads to neurodegeneration and plasticity that occurs along functional networks that depend on WM integrity [80, 81]. This is analogous to the putative mechanisms of hair cell loss and synaptopathy in presbycusis and hidden hearing loss. WM integrity follows a region-dependent, nonlinear trajectory throughout the lifespan, improving during development and declining after age 50 [82, 83]. Thus, the transmission and integration of vestibular and multi-sensorimotor information may be impacted differently in younger versus older adults. Together, these findings highlight how age-related changes in peripheral vestibular function coincide with evolving

WM structures, possibly forming the neurophysiological substrate for the motor and cognitive changes often observed in older adults. These considerations are important as older adults with vestibular dysfunction are at increased risk of balance and cognitive impairment [84]. Consequently, it is crucial to evaluate how these age-related WM changes translate into motor and cognitive outcomes, as discussed next.

### 4.2 Implications for Higher-Order Behaviors

Higher-order motor control and cognitive function both rely on vestibular input and WM microstructure. In the following subsections, we first examine how these WM findings may affect sensorimotor control in older adults (Section 4.2.1) and then explore potential cognitive ramifications (Section 4.2.2).

#### 4.2.1 Sensorimotor Control

Our findings highlight differential relationships between vestibular end-organ functions and the integrity of WM pathways involved in sensorimotor control in older adults. Higher canal and utricular function were associated with both lower and higher WM integrity against the backdrop of age-related declines. These findings are important, as several studies have shown that older adults with vestibular dysfunction have slower gait, longer and slower steps during normal speed walking, increased postural sway, altered head kinematics, and increased risk of falls [13, 85, 86, 87, 88, 84].

Higher canal function correlated with lower integrity in the left internal capsule, left sagittal stratum, the right column and body of the fornix, and left posterior thalamic radiation tract. These multi-sensorimotor areas in the longitudinal pathway connect the spinal cord, brainstem, basal ganglia, thalamus, and cerebral cortex. The sagittal stratum connects the occipital, temporal, and frontal lobes and transmits vestibular and visual information. Owing to the circuitry of these tracts, they play a role in motor planning and control of balance, posture, and (oculo-)motor functions [89, 90, 5]. In contrast, we observed higher integrity in interhemispheric and associative tracts, including the left inferior fronto-occipital fasciculus, left cingulum (hippocampus), the right cingulum (cingulate gyrus), the left and right external capsule, and the genu of the corpus callosum with respect to higher canal function. The left inferior fronto-occipital fasciculus connects the inferior occipital lobe (BAs 17, 18, 19, 37, 39) to the orbito-frontal cortex, superior, middle, and inferior frontal gyri (BAs 8, 10, 12, 45, 47, 9/46) via the ventral external capsule [91]. It is involved in speech production, semantic language processing, goal-oriented behavior, and visual switching tasks. Of note, structural changes in the corpus callosum have similarly been linked to motor coordination and balance problems in aging [92, 49] and, therefore, may be important for compensating or exacerbating vestibular-related motor impairments.

Higher utricular function was linked to lower integrity the right retrolenticular limb, possibly reflecting an age-related shift in reliance on vestibular signals. However, higher utricular function also correlated with higher integrity in the left column and body of the fornix and the right anterior limb of the internal capsule, suggesting preserved cognitive and motor relay pathways that may support memory function (e.g. verbal or visuospatial) and lateralized balance adjustments [46].

Overall, our findings suggest that that variations in vestibular end-organ function may selectively influence multisensory WM tracts involved in balance, postural stability, and motor planning in older adults. While these data highlight how vestibular deficits can reshape motor planning and balance pathways, there is increasing evidence that vestibular function also plays an important role in cognitive abilities. The next section discusses how these vestibular-associated WM changes may affect higher-level cognitive abilities.

#### 4.2.2 Cognitive Abilities and Cognitive-Sensorimotor Interactions

Several studies have highlighted the role of vestibular function in the trade-off between motor planning and execution and cognitive control. Vestibular patients must exert more cognitive control to maintain proper coordination during locomotion [93, 94]. It is thought that cognitive control over motor function and balance increases with aging [49].

Peripheral and central vestibular damage can lead to impairments in spatial orientation and self-motion perception. Traumatic brain injury accompanied by vestibular agnosia was linked to imbalance via damage to the right inferior longitudinal fasciculus [95]. Vestibular agnosia, in the presence of traumatic brain injury, disrupts connectivity in the right superior longitudinal fasciculus, left posterior thalamic radiation, bilateral superior corona radiata, and right posterior corona radiata and impacts self-motion processing, specifically in the yaw-plane—the plane of motion best sensed by the utricle and horizontal semi-circular canal [46]. Damage to the limbs of the internal capsule and to the thalamus have been associated with dizziness [96], rotational vertigo [97], and spatial neglect [2]. Canal dysfunction has been associated with dizziness and rotational vertigo, and utricular dysfunction has been associated with tilt illusions. It has been suggested that tilt illusion in Parkinson’s disease and spatial neglect result from damage to the limbs of the internal capsule and to the thalamus by failures of higher-order (vestibular) sensory integration [98, 2].

Yaw-plane motion transduced by the horizontal canal and utricle may play an important role in pathways integral for visual-vestibular coordination. The posterior thalamic radiation is directly connected to the sagittal stratum, which contains the medial longitudinal fasciculus and ipsilateral vestibulo-thalamic tract that transmits vestibular (e.g VOR) information to the lateral thalamic nuclei and in turn to the broader multi-sensorimotor vestibular network. This tract plays a role in the subjective visual vertical and is thought to convey signals for navigation, self-motion perception, and action [90, 5]. This is consistent with findings that thalamic multisensory integration is vital for stabilizing vision during head movements, an ability that often diminishes with age [90, 5]. Furthermore, these yaw motion pathways may be important for balance. Age-related horizontal canal dysfunction is associated with decreased performance on the standing on foam with eyes closed (FEC) balance test [99, 85]. Vestibular agnosia severity, postural instability on the FEC test, poorer VOR gain thresholds, and poorer vestibular-perceptual thresholds of yaw rotations have been linked to lower FA and higher MD (poorer WM integrity) in the inferior longitudinal fasciculus (part of the sagittal stratum) in the right temporal lobe [95].

The anterior limb of the internal capsule, the fornix, and the cingulum are important WM tracts in the Papez circuit which is important for memory and emotion function. The column and body of the fornix are major outputs to the mammillary bodies of the hypothalamus which project to the anterior nuclei of the thalamus–the home of the head-direction system. The anterior limb of the internal capsule carries thalamocingulate tract fibers which connect the anterior thalamic nuclei to the cingulate gyrus. The cingulum transmits sensory information from the cingulate gyrus and various association cortices via the parahippocampal gyrus, to the entorhinal cortex, in turn to the hippocampus. A growing body of literature suggests that vestibular dysfunction impacts the entorhinal cortex, hippocampus, and spatial cognitive ability [100, 17, 18]. Furthermore, as these tracts are involved in executive and visuospatial attention, control, and working memory, vestibular dysfunction may lead to their degradation which may explain the association with deficits in verbal initiation and digit span [101].

In summary, these possible lateralization patterns and associations with higher-order cognitive abilities suggest that age-related vestibular loss may not only impact the WM involved in motor control but also in visuospatial attention, executive function, and possibly real-world activities like driving.

### 4.3 Strengths and Limitations

We point out several strengths of this study. We used a large sample size of healthy, older adults who had prospective DTI scans and physiological assessments. To our knowledge, the BLSA is the only prospective study that offers both peripheral vestibular testing data and structural neuroimaging. We also implemented a robust and highly quality controlled analysis pipeline. Data was quality controlled by experts at the NIA. DTI scans were processed and segmented using a state-of-the-art pipeline. We avoided parametric assumptions by using permutation testing of our statistical hypotheses. Additionally, we achieved robust estimates of the spread of the effects of interest using accelerated and bias corrected bootstrapping. We also performed FDR-correction to account for multiple independent comparisons.

This study has limitations. Most of the observed associations did not survive FDR correction, though the goal of this first analysis of its kind was in hypothesis generation. The diffusion metrics used in this study are indirect measures of water diffusion in the brain, not direct measures of WM integrity. While we interpret our findings in terms of microstructural integrity, many well-known ambiguities arising from the underlying anatomy, such as fiber crossings and fiber density alterations, prevent clear interpretation [102, 103]. While several techniques, like HARDI or Qball imaging, have been developed to resolve such ambiguities, the true nature of vestibular-associated WM integrity effects should be derived from gold standard histology. Stemming from the limitations of classic diffusion imaging, the WM atlas definitions have ambiguities, resulting from inability to distinguish high granularity regions or certain portions of fiber tracts [89]. Furthermore, our diffusion metrics were averaged over entire WM labels which encompass many fibers and regions. We cannot describe more specific spatial relationships, as a result. With respect to the study sample, because the BLSA participants in this study were older (aged 60+), our results may not extend to younger subjects with vestibular dysfunction.

### 4.4 Future Work

There are several open questions that warrant further study. Whether vestibular loss accelerates the decline of WM integrity over time is unknown. If vestibular loss does accelerate WM decline, understanding the sequence of changes in each pathway will help to identify vulnerable networks for early intervention. Furthermore, future work will investigate whether WM mediates vestibular-associated deficits in motor and cognitive abilities.

## 5 Conclusion

This study is one of the first to demonstrate the differential effects of age-related peripheral vestibular function on structural integrity of WM pathways crucial for both motor and cognitive performance in healthy, older adults. Alterations to WM microstructure may explain the connection between vestibular dysfunction and the motor and cognitive abilities in older adults–preserving certain functions and curtailing others. Early identification of such neuroimaging biomarkers and targeted interventions may help mitigate the adverse effects of vestibular decline on healthy aging.

## Acknowledgments

This work was supported in part by the National Institute on Aging [grant number R01 AG057667, R01 AG073115], National Institute on Deafness and Other Communication Disorders [grant number R03 DC015583], and National Institute of Biomedical Imaging and Bioengineering [grant number P41-EB031771] and by the Intramural Research Program, National Institute on Aging, National Institutes of Health.

## Data Availability Statement

Data from the BLSA are available on request from the BLSA website (blsa.nih.gov). All requests are reviewed by the BLSA Data Sharing Proposal Review Committee and are also subject to approval from the NIH institutional review board.

## Code Availability Statement

The scripts used in this study can be shared by the corresponding author upon reasonable request.

